# Assessing the Role of Socioeconomic and Comorbid Factors on Nutritional Status Among Older Adults

**DOI:** 10.1101/2025.07.09.25331245

**Authors:** Mohoshina Karim, Partho Sharothi Das, Tasrima Trisha Ratna, Ishbat Ahmed, Mohima Sharmin, Syeda Saika Sarwar, Millat Hossain, Shahada Sharmin Mim, Joynal Abedin Imran, Shahriar Hasan, Marzana Afrooj Ria

## Abstract

Malnutrition in old age is a serious worldwide health concern, with complicated relationships between socioeconomic, health, and physiological factors. This research examines the influence of socioeconomic status, comorbidities, and sociodemographic factors on the nutritional status of older people in Bangladesh. The aim of this study is to evaluate the effect of factors like age, gender, education, income, marital status, financial independence, diabetes, hypertension, kidney disease, and cardiovascular disease on the risk of malnutrition among older people. A cross-sectional survey was performed with 622 participants at three of the biggest hospitals in Dhaka, Bangladesh. A validated self-reported questionnaire was utilized to obtain the data. Multinomial logistic regression was used to analyze the relationship between the explanatory factors and nutritional status, and Mini Nutritional Assessment (MNA) was used to evaluate nutritional status. The research indicated that older age (70 and older), female sex, illiteracy, lower income, financial dependence, and chronic disease conditions like diabetes, kidney disease, and cardiovascular disease were all strongly linked with a higher risk of malnutrition. Multinomial logistic regression indicated that those aged 70 and older were 7.25 times more likely to be malnourished than those in the 60-69-year-old age group. In the same way, participants with diabetes and with kidney disease had much increased chances of malnourishment. Malnutrition among older people is closely related to sociodemographic and comorbid factors. This paper emphasizes the imperatives of evidence-based interventions to enhance the nutritional status of at-risk subject populations, that is, older people with chronic conditions and lower socioeconomic status. Broader education, healthcare services, and social support are critical to reduce malnutrition risk in this group.

## Introduction

Nutrition is an important health factor in older adults, with far-reaching impacts on their physical, cognitive, and overall health. A proper diet plays an important role in maintaining muscle mass and metabolic health, both of which have strong associations with physical activity levels as well as skeletal muscle maintenance (1,2). However, most older adults experience nutrient deficiencies as a result of declining appetite, dietary limitations, as well as social isolation that contributes to adverse health outcomes such as frailty as well as cognitive decline (3,4).

Physical health in older adults’ benefits from nutrition. Research indicates a positive relationship between nutritional status and quality of life (QoL), and positive mental health status and reduced rates of depression in relation to improved eating patterns (5–7).

Yet, malnutrition in older populations is now an increasingly recognized worldwide health problem. Prevalence is highly variable across populations and settings, depending on geographic location, socioeconomic status, and assessment tool. In 2021, more than 97.6 million malnutrition cases were reported worldwide in older individuals, an ominous rise from 44.36 million in 1990 (8). The overall rate in worldwide prevalence has marginally lowered (−0.32% per year), but the absolute number has risen substantially, indicating increased scope for the condition (8).

In America, malnutrition has been responsible for a considerable number of fatalities in older populations, with 93,244 reported fatalities from 1999 to 2020. Mortality rates that were adjusted for age increased from 10.7 per 100,000 in 1999 to 25.0 per 100,000 in 2020, with an evident acceleration in rates after 2013 (9). The trend suggests an emerging public health problem of malnutrition in older adults even in high-resource environments.

In Singapore, 22.3% of community-dwelling older adults were found to be at moderate to high risk of malnutrition(10). Poor nutrition patterns were prevalent, including 90.5% of older adult Singaporeans exceeding suggested amounts for added sugars (10). Approximately 68.5% of men as well as 57.1% of women had sizable portions eating excessive amounts of saturated fats (10). This trend is clearly shown in a longitudinal study from China from 2008 to 2018. Underweight in older Chinese adults declined from 20.05% to 7.87%, overweight increased from 12.82% to 28.45%, and obesity went from 1.62% to 4.95% (11). This is part of wider global nutrition transitions related to urbanization, economic growth, and shifts in food systems.

Older adults’ malnutrition greatly inflates healthcare expenditure through increased use of resources. About 12.6% of older adults in the community in Spain were malnourished according to GLIM criteria. (12). The economic burden surpasses direct treatment costs. Older adults in the UK at risk of malnutrition (16.5% of those ≥50) lost 153,476 years of life lost (YLL) from premature death, with mortality rates at 755 per 100,000 person-years (13).

Older adult malnutrition is one of the biggest health challenges facing the world, closely linked to numerous socioeconomic factors. Age is one factor that is determined to increase with advanced years, which has been associated with higher malnutrition risk. Older adults between 60-65 years and above 75 years have been found to have high nutrition risk by studies (14). Another study in Mexico showed that malnutrition rises with older years, to 5% in people over 80 years (15).

Gender is also crucial in determining older adults’ malnutrition status, though gender’s relation to malnutrition differs between studies. Male were found to be predictive of a higher risk for malnutrition by the Hellenic Longitudinal Investigation (16). On the other hand, male were more likely to be malnourished than female in a South African-based study (17). Different results could explain variations in social and cultural factors between regions that impact nutrition.

Living arrangements and marital status also contribute to the nutritional status of older adults. Single, divorced, or widowed individuals have been found to have an increased risk for malnutrition in comparison to those who are married (16,18). This trend could indicate that social care and spousal support might have a protective effect on nutrition.

Education is another socioeconomic factor that is relevant to determining older adults’ nutrition. The Hellenic Longitudinal Study of Aging and Diet also demonstrated that lower education was strongly correlated with a higher risk of malnutrition in urban older adults (16). This educational gradient in risk for malnutrition is also attested to by evidence from India, where older people from less-educated backgrounds were more likely to have been found to have underweight status (19). Education is thus significant in tackling malnutrition in older populations.

Economic resources essentially play a crucial role in determining an older adult’s capability to acquire sufficient nutrition. Several studies have found that low socioeconomic status is an independent predictor for malnutrition risk. In one cross-sectional observational study in Greece, researchers determined that older adults with lower socioeconomic status possessed higher risks for malnutrition (16). In another study in India, older adults with per-month household income ≤R1600 per month (about 133 USD) were at higher risks for being malnourished or being at risk for malnutrition (17).

Financial vulnerability enhances malnutrition risk by restricting access to nutrient-rich foods. In India, older adults with limited monthly incomes experienced 20-40% increased risks for undernutrition than their higher-income counterparts (20). Unemployment and pension benefits also intensified this risk, especially among farm workers, who experienced dual risks of being undernourished and food insecure(20).

Diabetes mellitus directly impacts malnutrition risk in older populations via intricate biological, socioeconomic, and clinical mechanisms. Diabetes duration for more than 10 years even triples (OR=2.99) the risk of malnutrition (21). In addition, over 51.1% of older individuals with type 2 diabetes have sarcopenia ranging from possible to severe, promoted by mechanisms including insulin resistance, chronic inflammation, and inadequate nutrition (22).

Chronic kidney disease (CKD) is a serious health challenge in older adults with far-reaching consequences for nutrition. Evidence from studies indicated that malnutrition and sarcopenia have been most evident in older hemodialysis (HD) patients with rates significantly higher in comparison to non-HD older adults (23). Utilizing one systematic review involving the utilization of the Malnutrition-Inflammation Score (MIS), researchers noted a staggering 52.2% prevalence of malnutrition and inflammation among older hemodialysis patients (24).

Cardiovascular disease (CVD) is also an important contributor to malnutrition risk in older adults. A bidirectional relationship between malnutrition and CVD is suggested by recent studies. In a study among older Japanese adults with severe aortic stenosis who were to undergo transcatheter aortic valve implantation, 89.3% were at least mildly malnourished on one or more scores used in nutritional assessment (25). Also, trials employing the use of the Geriatric Nutritional Risk Index (GNRI) have shown that it is useful for forecasting cardiovascular mortality across diverse populations (26,27). In older adults with diabetes, malnutrition, as quantifiable by the GNRI, was strongly related to all-cause mortality as well as to mortality from cardiovascular disease (27).

Geriatric malnutrition is an emerging health dilemma across the world that affects physical and cognitive functionality. Reduced appetite, social isolation, diabetes, and cardiovascular disease all raise malnutrition risk. Low income and education further complicate this. This study seeks to determine sociodemographic status and comorbidity factors that contribute to malnutrition in older individuals in order to inform evidence-based targeted interventions that can improve their nutrition.

Geriatric malnutrition is an expanding problem and serious health issue worldwide, with tremendous consequences for physical functioning in elderly adults. It is crucial to identify specific sociodemographic and comorbid factors contributing to malnourishment prevalence in older adults, as it strongly influences health outcomes. Based on this identification, this research intends to provide evidence-based targeted interventions that would effectively enhance the nutritional health of older adults. Prevention and management of geriatric malnutrition will enhance healthy aging, enhance the quality of life, and reduce healthcare costs due to complications.

## Materials & Method

### Study population

This study was a cross-sectional and carried out in three major hospitals. The name of the hospitals was, Dhaka Medical College Hospital (DMCH), National Institute of Traumatology and Orthopedic Rehabilitation (NITOR), and Bangladesh Institute of Research and Rehabilitation in Diabetes, Endocrine and Metabolic Disorders (BIRDEM). These are three major health institutions in Dhaka, Bangladesh, with diversified patient populations offering specialized medical care. The study was carried out from 20 April 2024 to 29 December 2024.

### Sampling process

There were 622 elderly participants in total. The participants were sampled through simple random sampling. Participants with written consent were enrolled. Whenever a participant refused, the next available eligible participant in line was approached. This helped avoid bias in sampling and reduced non-response bias, enhancing the validity of the study.

### Survey instruments

The data were collected using the English interviewer-administered validated questionnaire by face-to-face interviews with the participants in the studies. The participants were informed of the research purpose prior to the questionnaire and were informed that they were participating voluntarily. The questionnaires collected socio-demographic details and determined nutrition status.

### Data analysis

Data were initially handled in an MS Excel sheet, after which Stata version 16 was utilized for analysis. The demographic details were summarized from data using descriptive statistics for comorbidities as well as frequency for nutritional status. Multinomial logistic regression was applied for determining association between multiple socio-demographic and comorbid variables (such as age, gender, marital status, education, monthly family income, financial independence, diabetes, kidney disease, cardiovascular disease, and hypertension) with nutritional status for analytical purposes. Adjusted as well as unadjusted regression were carried out for individual explanatory variables.

### Socio-demographic and Comorbidity related characteristics

The explanatory variables in this study are categorized as follows: Age group (60 to 69 years vs 70 and above years), Gender (Male vs Female), Marital Status (Married vs Unmarried), Education (Illiterate vs Primary vs Secondary and above), Monthly family income (25,000 and below, 25,001 to 49,999, 50,000 and above), Financial independence (Independent vs Dependent), Diabetes (Present vs Absent), Hypertension (Present vs Absent), Kidney disease (Present vs Absent), and Cardiovascular disease (Present vs Absent).

### Outcome Measures

The main outcome measure is Nutritional Status, for which the Mini Nutritional Assessment (MNA) tool (28). MNA tool allocates participants into three categories based on the scores that range from normal nutrition status (MNA score of 24 or above), at risk of malnutrition (MNA score of 17-23.5), and Malnourished (MNA score below 17) (28).

For validity purposes, forward and backward translation and field tests were carried out with 40 elderly subjects belonging to the same population. The instrument’s reliability was established by a Cronbach’s alpha value of 0.72, reflecting acceptable internal consistency.

### Ethical consideration

Ethical clearance was obtained from the Institutional Review Board of the National Institute of Traumatology and Orthopedic Rehabilitation (NITOR/PT/93/lRB/2024/02). Informed consent was signed by all participants before data was collected.

## Result

Figure 1 illustrates that 30.1% of the participants were of normal nutritional status, 58.7% at the risk of malnutrition, and 11.3% were malnourished.

**Fig. 1.**
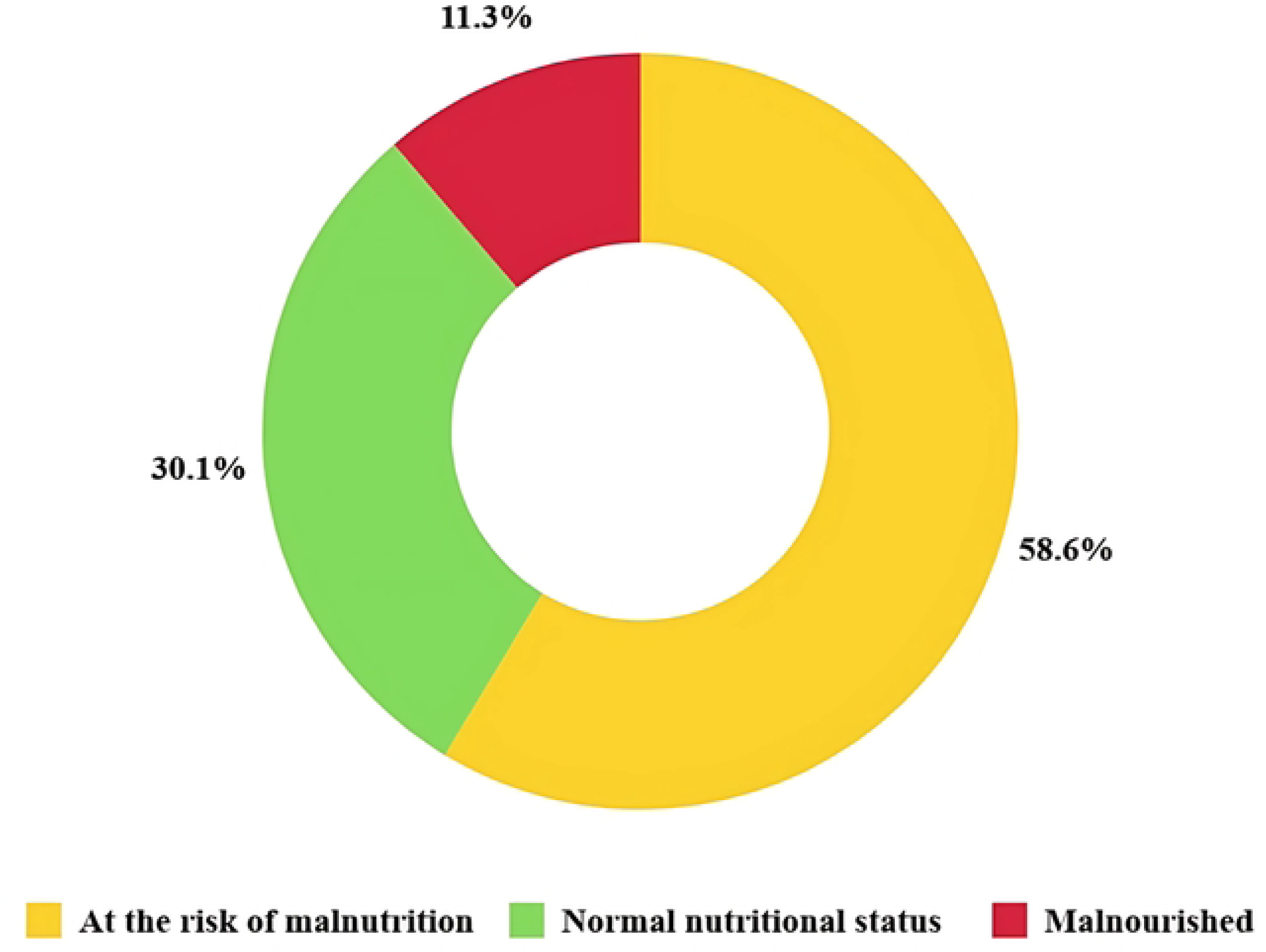
Proportion of nutritional status among the participants (n = 622)

Based on this study, we have noted that, individuals who were 70 years old and older were more likely to be malnourished (26.9%) in comparison to those 60-69 years old had mostly normal nutritional status (32.7%) (Table 1). Women are more likely to be malnourished (13.4%) in comparison to men (7.7%). Married participants tend to be better off in terms of being normally nourished (34.6%) and less likely to be malnourished, but the unmarried have a greater proportion to be malnourished (18.7%).

**Table 1.**
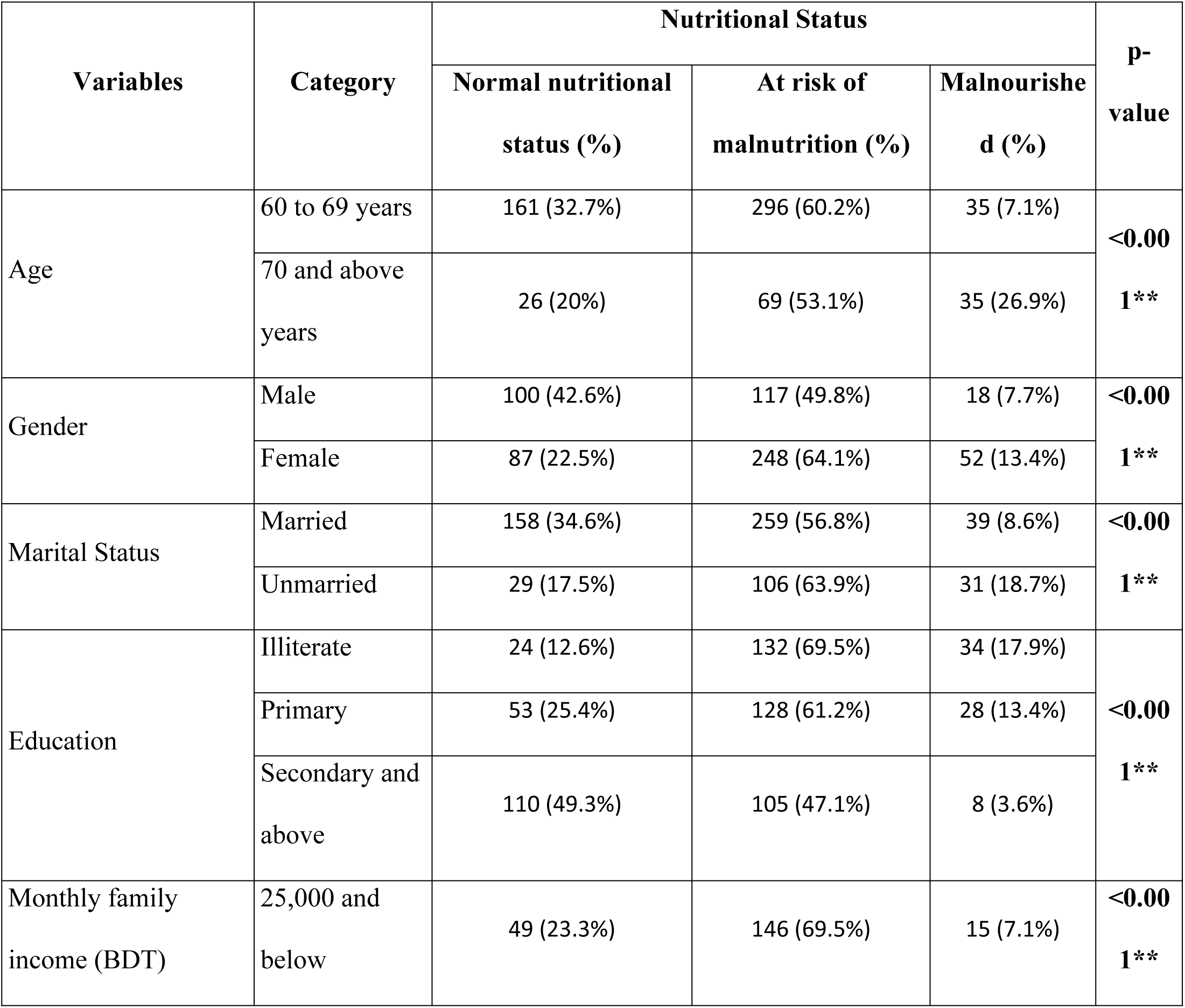
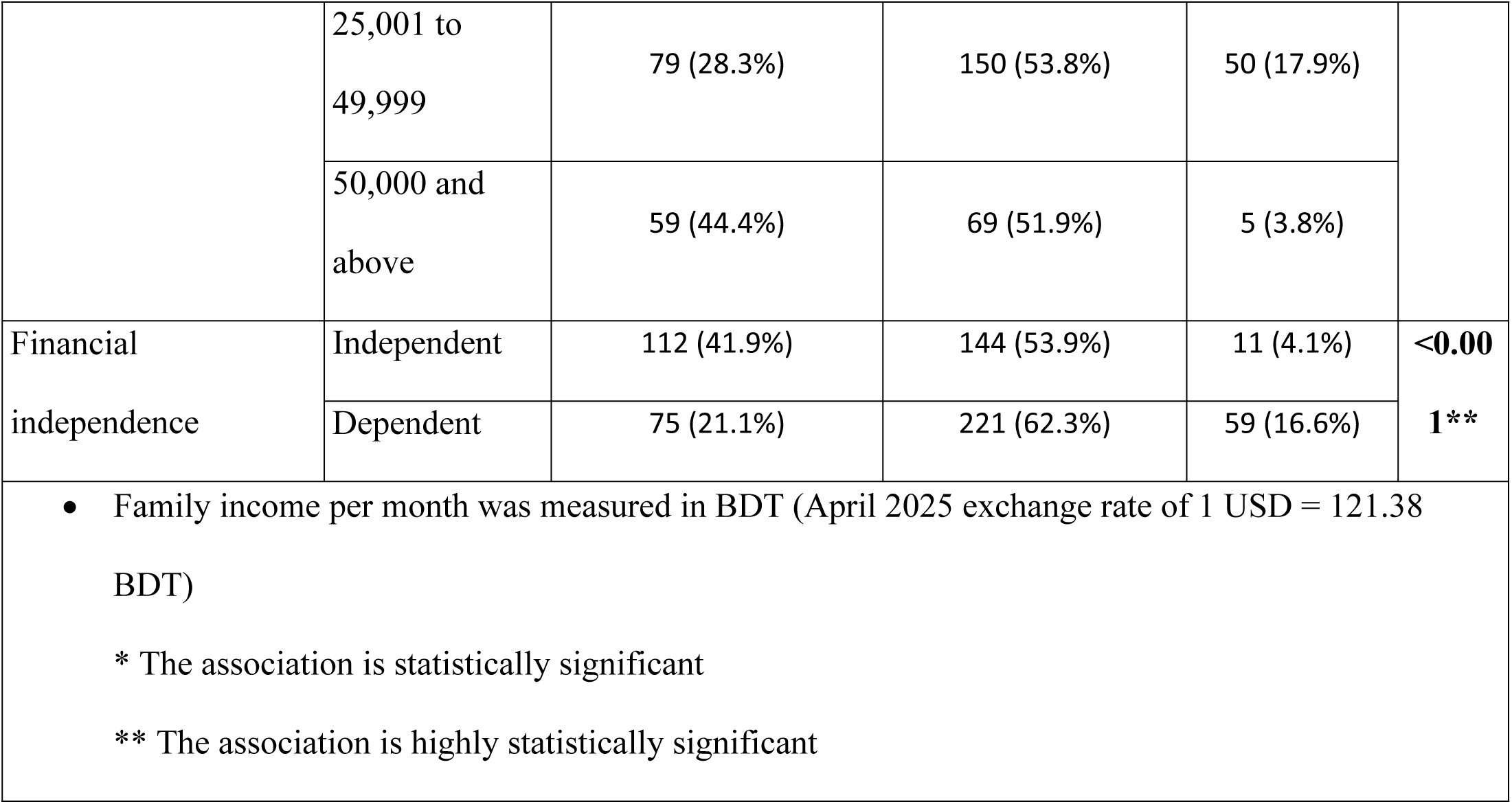
Association of Sociodemographic status with Mini Nutritional Score (*n* = 622)

Illiterate participants have a high proportion at risk of being malnutrition (69.5%). Those in income range 25,001 to 49,999 BDT have a relatively higher proportion of malnourished individuals (17.9%) from that in the highest income group (7.1%). Participants who were financially dependent, is associated with higher proportions being malnourished (16.6%), compared to independent participants where 4.1% are malnourished. From table 2 we observed that Individuals who have diabetes, have higher proportions being malnourished (24.4%), compared to non-diabetic individuals (4.6%).

**Table 2.**
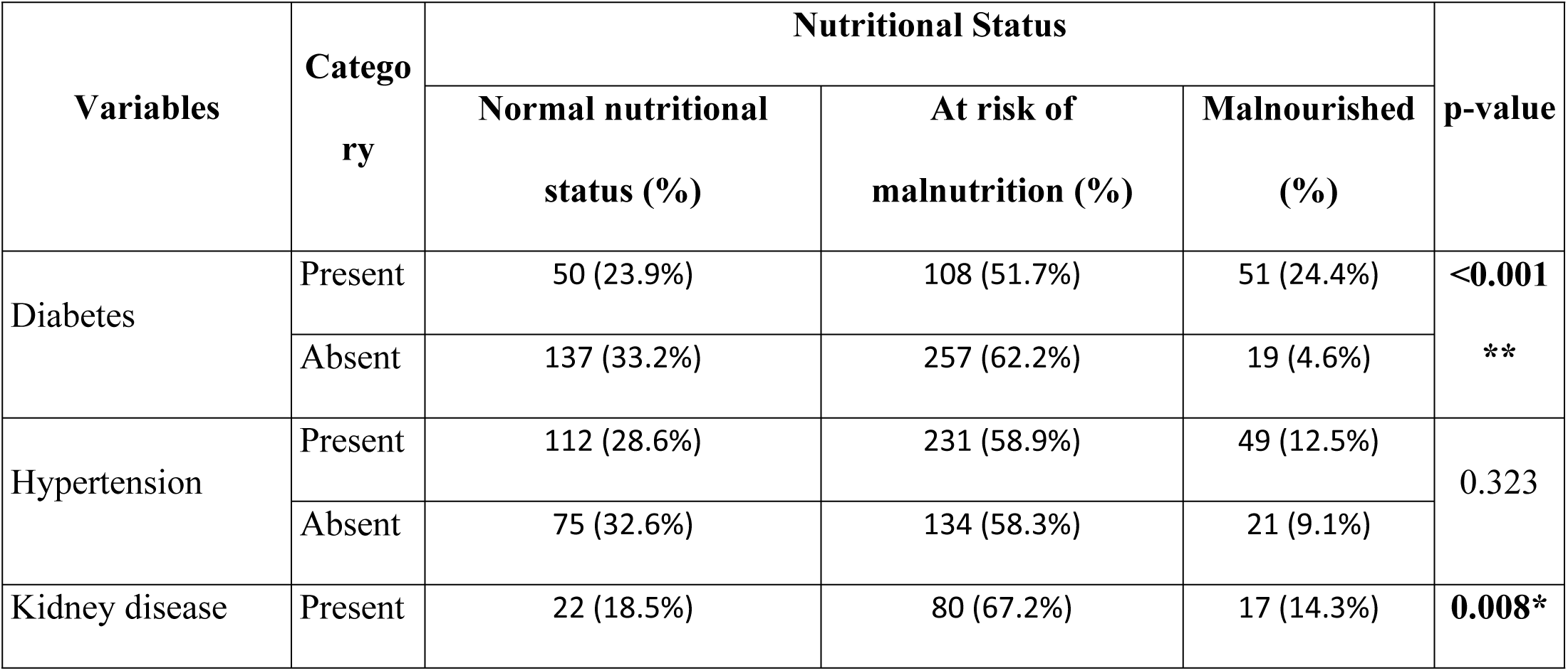
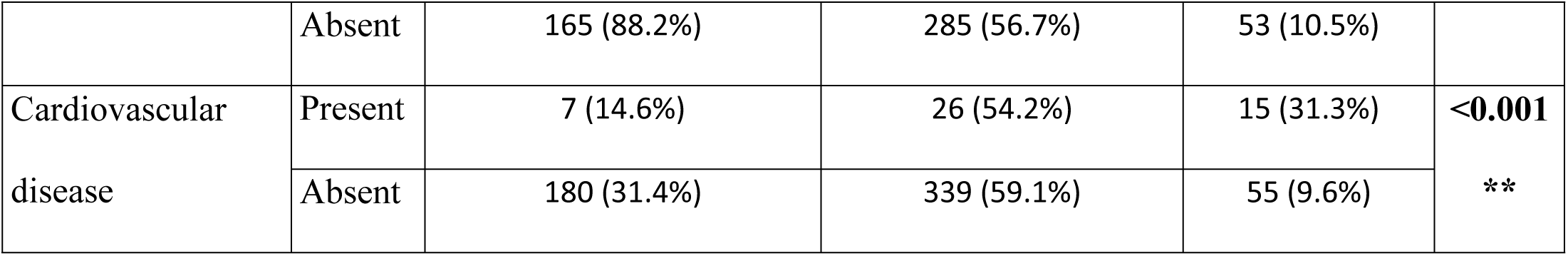
Association of comorbid factors with Mini Nutritional Score (*n* = 622)

Hypertension is linked to a higher proportion of being malnourished individuals (12.5%) compared to non-hypertensive individuals. Individuals with kidney disease have a higher proportion of malnourished individuals (9.1%) compared to non-kidney disease individuals. The proportion of malnourished is considerably higher in individuals with cardiovascular disease (31.3%) as compared to non-cardiovascular disease individuals (9.6%).

Furthermore, nutritional status was significantly associated with age (p < 0.001), gender (p < 0.001), marital status (p < 0.001), education (p < 0.001), monthly family income (p < 0.001), financial independence (p < 0.001), diabetes (p < 0.001), kidney disease (p = 0.008), cardiovascular disease (p < 0.001) (table 1 and 2).

In table 3, respondents who were 70 years and older were 7.25 times more likely to be malnourished (AOR = 7.25; 95% CI: 3.10–16.95, p < 0.001) than respondents aged between 60 to 69 years. Female respondents were 1.89 times more likely to have malnutrition risk (AOR = 1.89; 95% CI: 1.15–3.12, p = 0.012) compared to male.

**Table 3:** Logistic regression analysis of Sociodemographic status and comorbid factors with Mini Nutritional Score (Multinomial Logistic regression)

| Variables | Category | Nutritional Status |  |  |  |  |  |  |  |
| --- | --- | --- | --- | --- | --- | --- | --- | --- | --- |
|  |  | Normal nutrition vs At risk of malnutrition |  |  |  | Normal nutrition vs Malnourished |  |  |  |
|  |  | Unadjusted model |  | Adjusted model |  | Unadjusted model |  | Adjusted model |  |
|  |  | OR<br>95% CI | p-value | OR<br>95% CI | p-value | OR<br>95% CI | p-value | OR<br>95% CI | p-value |
| Age | 60 to 69 years | Reference |  |  |  | Reference |  |  |  |
|  | 70 and above years | 1.44<br>(0.88-2.35) | 0.142 | 1.60<br>(0.90-2.83) | 0.105 | 6.19 (3.31<br>-11.57) | <0.001** | 7.25<br>(3.10-16.95) | <0.001** |
| Gender | Male | Reference |  |  |  | Reference |  |  |  |
|  | Female | 2.43<br>(1.69-3.49) | <0.001** | 1.89<br>(1.15-3.12) | 0.012* | 3.32<br>(1.80-6.10) | <0.001** | 2.18<br>(0.79-5.97) | 0.129 |
| Marital Status | Married | Reference |  |  |  | Reference |  |  |  |
|  | Unmarried | 2.22<br>(1.41-3.51) | <0.001** | 1.06<br>(0.60-1.86) | 0.828 | 4.33<br>(2.33-8.01) | <0.001** | 1.72<br>(0.74-4.02) | 0.204 |
| Education | Secondary and above | Reference |  |  |  | Reference |  |  |  |
|  | Primary | 2.53<br>(1.66-3.83) | <0.001** | 2.08<br>(1.32-3.28) | 0.001* | 7.26<br>(3.10-17.01) | <0.001** | 4.97<br>(1.77-13.95) | 0.002* |
|  | Illiterate | 5.76<br>(3.45-9.60) | <0.001** | 3.93<br>(2.24-6.90) | <0.001** | 19.47<br>(8.01-47.32) | <0.001** | 13.08<br>(4.40-38.82) | <0.001** |
| Monthly family income (BDT) | 50,000 and above | Reference |  |  |  | Reference |  |  |  |
|  | 25,001 to 49,999 | 1.62<br>(1.04-2.52) | 0.110 | 1.88<br>(1.12-3.13) | 0.015 | 7.46<br>(2.80-19.88) | <0.001** | 13.18<br>(3.97-43.72) | <0.001** |
|  | 25,000 and below | 2.54<br>(1.58-4.09) | <0.001** | 2.44<br>(1.39-4.30) | 0.002* | 3.61<br>(1.22-10.64) | 0.020 | 5.46<br>(1.46-20.36) | 0.011* |
| Financial independence | Independent/Partially independent | Reference |  |  |  | Reference |  |  |  |
|  |  | Normal nutrition vs At risk of malnutrition |  |  |  | Normal nutrition vs Malnourished |  |  |  |
|  |  | Unadjusted model |  | Adjusted model |  | Unadjusted model |  | Adjusted model |  |
|  |  | OR<br>95% CI | p-value | OR<br>95% CI | p-value | OR<br>95% CI | p-value | OR<br>95% CI | p-value |
|  | Dependent | 2.29<br>(1.59-3.28) | **<0.001\*\*** | 1.79<br>(1.15-2.79) | 0.010* | 8.00<br>(3.94-16.24) | **<0.001\*\*** | 3.67<br>(1.53-8.80) | **0.004\*** |
| Diabetes | Absent | Reference |  |  |  | Reference |  |  |  |
|  | Present | 1.15<br>(0.77-1.70) | 0.483 | 0.97<br>(0.61-1.52) | 0.901 | 7.35<br>(3.96-13.64) | **<0.001\*\*** | 5.17<br>(2.42-11.01) | **<0.001\*\*** |
| Kidney disease | Absent | Reference |  |  |  | Reference |  |  |  |
|  | Present | 2.1 (1.26-3.50) | **0.004\*** | 2.20<br>(1.25-3.87) | **0.006\*** | 2.40<br>(1.18-4.86) | **0.015\*** | 4.09<br>(1.62-10.35) | **0.003\*** |
| Cardiovascular disease | Absent | Reference |  |  |  | Reference |  |  |  |
|  | Present | 1.97<br>(0.83-4.63) | 0.119 | 1.30<br>(0.51-3.26) | 0.575 | 7.01<br>(2.72-18.07) | **<0.001\*\*** | 2.66<br>(0.83-8.56) | 0.093 |
| Hypertension | Absent | Reference |  |  |  | Reference |  |  |  |
|  | Present | 1.15<br>(0.80-1.65) | 0.437 | 1.06<br>(0.71-1.60) | 0.748 | 1.56<br>(0.86-2.81) | 0.137 | 1.01<br>(0.49-2.09) | 0.967 |

Illiterate respondents were 13.08 times (AOR = 13.08; 95% CI: 4.40-38.82, p < 0.001) and primary educated respondents were 4.97 times (AOR = 4.97; 95% CI: 1.77–13.95, p = 0.002) more likely to be malnourished. The respondents with monthly income between 25,001 and 49,999 BDT were 13.18 times (AOR = 13.18 95% CI: 3.97–43.72, p < 0.001) and monthly income of 25,000 BDT and below were 5.46 times (AOR = 5.46; 95% CI: 1.46–20.36, p = 0.011) more likely to experience severe malnutrition. The financially dependent respondent were 3.67 times more likely to suffer from severe malnutrition (AOR = 3.67; 95% CI: 1.53–8.80, p = 0.004).

The people with diabetes were 5.17 times more likely to be malnourished (AOR = 5.17; 95% CI: 2.42–11.01, p < 0.001). Individuals with kidney disease were 4.09 times more likely to be malnourished (AOR = 4.09; 95% CI: 1.62–10.35, p = 0.003).

No multicollinearity between the independent factors was seen. The regression model showed that variation in the dependent variable can be explained by the independent variables (age, gender, education, monthly income, financial independence, diabetes, and kidney disease) in the model.

## Discussion

Malnutrition is one of the major health challenges for the aging people of the world, with complex causes across physiologic, socioeconomic, functional, and environmental areas. Studies have shown that older adults have specific challenges in nutrition that make them at considerably an increased risk for developing malnutrition compared to younger populations. The prevalence of older adults at the risk of malnutrition varies between 13.5% and 58.2% among various populations and environments, and socially isolated and institutionalized older adults encounter higher risks (29–31).

This study found that respondents 70 years old and older tend to have higher malnutrition rates (26.9%) in comparison to respondents between 60-69 years with odds of 7.25 times. This is in line with evidence suggesting that malnutrition risk rises with advanced age, with older adults 75 years old and older having malnutrition rates 3.81 times higher than for 65-74 years (32).

Aging is associated with dramatic changes in body composition, which directly influence nutrition. As one grows older, there is a natural reduction in strength and muscle mass (sarcopenia), bone density loss, and higher fat mass (33,34). In addition, changes in taste and smell with aging influence nutrient consumption by older adults. Decreased taste and smell sensitivity decrease consumption enjoyment and appetite for food, which in some cases may decrease food consumption (33). Older adults on the other hand often have changes in digestion such as less stomach acid production, slower gastric emptying, and alterations in bowel anatomy (35).

Older adults have evident gender-specific trends in their nutritional status based on biological, social, and environmental variables. Though malnutrition rates don’t favor one gender equally across all studies, gender distinctions are apparent in risk factors, physiological effects, and health outcomes for older men compared to women.

Based on this study’s findings, we have noted that older females tend to have 1.89 times more chance to be malnourished (64.1%) in comparison to males (49.8%). This result concurs with another work when they mentioned that the odds in regard to being in a state of malnutrition were 2.46 times higher among women than among old men (32). However, in one study in Saudi Arabia, their finding was not similar. The older men had higher malnutrition prevalence by just 7% compared to women by 5% (36). On the other hand, one Finnish study among home-dwelling 75-year-old individuals revealed that malnutrition frequency was not statistically significantly disparate between sexes which was 12.7% on women and 8.6% on me (37). Such variations could possibly indicate varying social support systems, differing dietary habits, or differential accessibility to healthcare (36).

Married people were found in this study to have better usual nutrition status (34.6%) and were less likely to be malnourished, but for unmarried, there is higher proportion to be malnourished (18.7%). Another study revealed that unmarried is related to higher malnutrition risk vs. being married (16). In cross-sectional study from Wuhan, China, marital status was one of the best predictors for nutrition status in community-dwelling older individuals, in addition to the number of chronic diseases, age, and functional status (38).

Marriage’s protective role seemingly is complex. Married people derive benefits from routine social contact as well as caregiving in reciprocity, which might have positive impacts on dietary consumption (39). Married couples tend to have greater incomes in their households that might allow for improved food security as well as availability of nutritious foods (40).

In this research, we found that Illiterate respondents were 13.08 times more likely to have chances to be malnourished. This is similar to another study on India and this one found close to 14.8 times higher risk of malnutrition for illiterate older people in comparison to their counterparts who were not illiterate, reflecting very high association in this group (41).

In contrast to the overwhelming evidence for an association between malnutrition and illiteracy, not all studies have concluded as such. In one study in Turkey among older adults residing in communities, education groups were not found to have any difference in malnutrition rates (42). This conflicting finding implies that context-specific variations in factors such as cultural background, social support systems, or local-level economic conditions may impact the association.

This study overwhelmingly shows that those with lower incomes have 13.18 times higher malnutrition rates compared to their higher-income counterparts. A sweeping study also showed that older adults from low-income groups have far greater malnutrition risks compared to their higher-income counterparts. A Japanese study comparing albumin concentrations in 6,528 older adults revealed that albumin concentrations were statistically significantly lower in those from low-income groups (−0.17 g/L) compared to their counterparts from middle-income groups (43). Nigerian studies found 74.9% of older adults from low-income rural areas had substandard dietary habits, 56.2% being overweight/obese which is a dual burdens interlinking poverty with undernutrition as well as metabolic diseases (44).

Emerging evidence also indicated complex relationships between diabetes mellitus and malnutrition risk in older populations, where diabetic elderly individuals experienced unique nutrition challenges often surpassing those experienced by their non-diabetic counterparts. In this study, we discovered that diabetes people were 5.17 times at risk for malnutrition compared to non-diabetic people. This is similar to a study in Northeastern Brazil in which diabetic patients were 3.7 more likely to have malnutrition (45).

Elderly CKD individuals have undue risks of malnutrition in comparison to their non-CKD counterparts that is influenced by disease-related metabolic derangement, strict dietary needs, and accelerated aging (46).This study found that individuals with kidney disease were 4.09 times more likely to be malnourished, which is in line with another study that reported 68% of older CKD individuals have low dietary protein consumption which results in increased malnutrition and frailty over those with usual dietary protein consumption (47).

## Conclusion

This study underscores the significant impact of socioeconomic factors and comorbidities on the nutritional status of older adults in Bangladesh. Key findings indicate that advanced age, female gender, low income, low education, financial dependence, and chronic conditions such as diabetes, kidney disease are strongly linked to an increased risk of malnutrition. These results highlight the urgent need for targeted interventions to address the nutritional needs of older adults, especially those facing multiple health and socioeconomic challenges. While this study provides valuable insights, further research is needed to establish causal relationships and evaluate the effectiveness of specific interventions. The findings of this research also have a limited generalizability because the population was drawn only from participnats from three major hospitals in Dhaka. In conclusion, addressing malnutrition in older adults requires a comprehensive approach focusing on both health and socioeconomic factors to improve their quality of life and reduce healthcare costs.

## Data Availability

All relevant data are within the manuscript and its Supporting Information files.

## Supporting information

S1 Appendix. Informed consent form. The written consent form was administered to the participants.

S2 Appendix. Questionnaire. The included questions comprised the questionnaire administered to the participants.

S3 Dataset. The raw data supporting the article.

## Acknowledgment

The authors extend thanks to the participants from Dhaka Medical College Hospital (DMCH), the National Institute of Traumatology and Orthopedic Rehabilitation (NITOR), and the Bangladesh Institute of Research and Rehabilitation in Diabetes, Endocrine and Metabolic Disorders (BIRDEM) for all the help and cooperation they extended throughout the research.

## Funding Statement

No funding was received from any public, commercial, or not-for-profit agency for the conduct of the study.

## Author Contribution

Conceptualization: Dr. Mohoshina Karim, Dr. Partho Sharothi Das, Dr. Mohima Sharmin, Shahriar Hasan

Data Curation: Dr. Partho Sharothi Das, Syeda Saika Sarwar, Shahada Sharmin Mim

Formal Analysis: Dr. Partho Sharothi Das, Tasrima Trisha Ratna, Shahriar Hasan

Investigation: Dr. Mohoshina Karim, Shahriar Hasan, Dr. Partho Sharothi Das, Joynal Abedin Imran,

Methodology: Dr. Mohoshina Karim, Dr. Partho Sharothi Das, Ishbat Ahmed, Joynal Abedin Imran

Project Administration: Dr. Mohoshina Karim, Dr. Partho Sharothi Das

Resources: Dr. Mohoshina Karim, Shahriar Hasan

Software: Tasrima Trisha Ratna, Ishbat Ahmed, Millat Hossain, Marzana Afrooj Ria

Supervision: Dr. Mohoshina Karim, Dr. Mohima Sharmin

Validation: Tasrima Trisha Ratna, Shahada Sharmin Mim, Dr. Mohima Sharmin

Visualization: Syeda Saika Sarwar, Millat Hossain, Joynal Abedin Imran

Writing – Original Draft: Dr. Partho Sharothi Das, Tasrima Trisha Ratna, Shahada Sharmin Mim

Writing – Review & Editing: Dr. Mohoshina Karim, Ishbat Ahmed, Syeda Saika Sarwar

## Declaration of Conflict of interest

The authors disclose that they have no competing financial interests or personal relations that might have appeared to have an effect on the work described in this paper.

## Notes

### Competing Interest Statement

The authors have declared no competing interest.

### Funding Statement

The author(s) received no specific funding for this work.

### Author Declarations

1. Institutional review board: National Institute of Traumatology and Orthopaedic Rehabilitation (NITOR) 2. Approval number: NITOR/PT/93/lRB/2024/02 3. Written consent was obtained

